# Trajectory of Hyper IgE complications

**DOI:** 10.64898/2026.09.11.26362846

**Authors:** Asteria Mao, Rui Xiao, Kathleen E. Sullivan, Ramsay Fuleihan, Charlotte Cunningham-Rundles, Jennifer Puck, Rebecca Marsh, Roshini S. Abraham, Luigi Notarangelo, Vaibhavi Vichare, USIDNET Network members

## Abstract

**Objective:** Our objective was to define the natural history of hyper IgE syndrome. Well, there have been a number of large cross-sectional studies. It has been difficult to provide anticipatory care for the growing adult population.

**Methods:** The USIDNET registry was used to identify patients with hyper IgE syndrome. We evaluated all patients regardless of genetic etiology. Complications were extracted as text from the problem list or ICD codes, and each complication was date stamped, which allowed us to estimate the age at which the complication arose.

**Results:** Many of the complications previously identified were seen in this cohort. The most feared complications were uncommon, however, developmental delay and psychosis affected nearly a fifth of the cohort which has not been previously reported.

**Conclusions:** As the number of adults with hyper IgE syndrome increases our awareness of late onset complications is important. Aspergillus and pneumatoceles were less common than seen in some other cohorts. However, the significant rate of psychosis will represent a significant challenge for clinicians.

## Introduction

The hyper IgE syndromes are Inborn Errors of Immunity that collectively are characterized by an elevated serum IgE level, eczema, and recurrent infections ^1^. Initial publications referred to it as Job’s syndrome and emphasized recurrent staphylococcal abscesses that lacked the classic signs of inflammation such as redness and tenderness ^2^. In the 1990s, recognition that the syndrome could have skeletal complications as well as dental anomalies simultaneously identified multiple kindreds with an apparent autosomal dominant inheritance ^3-5^. Today, it is recognized as a classic multisystem disorder and multiple genetic etiologies have been identified ^6, 7^. The most common form and the best characterized is due to autosomal dominant variants in the *STAT3* gene ^8, 9^. Most of the other forms of hyper IgE syndrome are due to genetic variants in the same pathway as STAT3. DOCK8 deficiency is an autosomal recessive condition, which was originally classified as one of the hyper IgE syndrome due to its prominent eosinophilia and elevated IGE. However, overtime, most of the complications have come to be recognized as a component of a T cell deficiency and it is now better classified as an actinopathy.

For decades, the management of patients with hyper IgE syndrome has been focused on the prevention and treatment of infections ^9, 10^. Some have been treated with immunoglobulin replacement due to a recognition that their IgG level may be normal but their capacity to make antibody responses to vaccines is compromised. In spite of that, patients develop pulmonary pneumatoceles and subsequent overgrowth of the cavities with aspergillus ^11^. Newer treatments with dupilumab and even bone marrow transplantation have been reported, but outcome studies are lacking and a major impediment is the lack of natural history reports ^12, 13^. The study was undertaken to better define the lifetime trajectory of hyper IgE syndrome using data from the USIDNET registry.

## Methods

87 unique patients carrying a diagnosis of hyper IGE syndrome identified in the USIDNET registry in 2026. We further extracted demographic characteristics, gene names, and complications. The USIDNET registry times stamps each entry with the days-of-life of the patient. From these data, we were able to bin the findings according to approximate ages. Data are displayed as both patient counts and event counts.

## Results

The demographic characteristics of the 87 unique patients are shown in Table 1. The sex and racial characteristics are typical for the registry. We then went on to collect the most frequent ICD diagnoses for each patient. As expected because it is part of the core clinical features for hyper IgE syndrome, eczema, pneumonia, and abscesses were common.

**Table 1.**

| Characteristic | N = 87<sup>1</sup> |
| --- | --- |
| Age (years) | 21.5 (13.0) |
| Sex |  |
| Female | 41 (47.1%) |
| Male | 46 (52.9%) |
| Race |  |
| Asian | 5 (5.7%) |
| Black or African American | 20 (23.0%) |
| Multiple Races | 4 (4.6%) |
| Other | 6 (6.9%) |
| Unknown | 3 (3.4%) |
| White | 49 (56.3%) |
| Ethnicity |  |
| Hispanic or Latino | 22 (25.3%) |
| Non-Hispanic or Non-Latino | 62 (71.3%) |
| Unknown | 3 (3.4%) |
| <sup>1</sup> Mean (SD); n (%) |  |

Candida has been increasingly recognized, particularly and infancy. Indeed, a quarter of the cohort carried a diagnosis of Candida. Unexpectedly we found a surprisingly high rate of psychosis and developmental delay in this cohort. This has not been previously reported for hyper IgE syndrome. Table 2 reports the most common diagnoses across the cohort as unique patients who carried those features.

**Table 2.**

| diagnosis_group | n_patients | percent |
| --- | --- | --- |
| Eczema | 58 | 66.67 |
| Pneumonia | 38 | 43.68 |
| Fracture | 31 | 35.63 |
| Abscess skin | 24 | 27.59 |
| Candida | 22 | 25.29 |
| Psychosis | 18 | 20.69 |
| Malignancy | 17 | 19.54 |
| Developmental delay | 16 | 18.39 |
| Scoliosis | 14 | 16.09 |
| Aspergillus | 8 | 9.20 |
| Pneumatocele | 5 | 5.75 |
| Mycobacteria | 4 | 4.60 |

To better understand the patient experience as an evolution of features, we identified the age at first diagnosis for the most common features identified in Table 3. We also performed an event count per patient, recognizing that some patients appear to have recurrent complications. Figure 1 displays the mean number of events per patient.

**Table 3.**

| diagnosis_group | n | mean_age | sd_age | median_age | Q1 | Q3 | min_age | max_age |
| --- | --- | --- | --- | --- | --- | --- | --- | --- |
| Abscess skin | 24 | 13.24 | 8.44 | 14.66 | 5.2 | 19.94 | 0.57 | 28.43 |
| Aspergillus | 8 | 28.02 | 17.01 | 24.98 | 20.86 | 31.16 | 5.62 | 60.33 |
| Candida | 22 | 9.93 | 8.87 | 7.39 | 1.89 | 17.39 | 0.41 | 24.92 |
| Developmental delay | 16 | 3.33 | 2.65 | 2.06 | 1.95 | 4.28 | 0.23 | 9.68 |
| Eczema | 58 | 9.06 | 10.4 | 4.31 | 1.76 | 14.81 | 0.05 | 60.62 |
| Fracture | 31 | 13.2 | 13.29 | 9.5 | 5.53 | 15.34 | 0.21 | 66.04 |
| Malignancy | 17 | 22.54 | 14.46 | 22.36 | 14.83 | 24.96 | 2.14 | 61.52 |
| Mycobacteria | 4 | 36.48 | 29.28 | 38.28 | 13.68 | 61.08 | 6.69 | 62.69 |
| Pneumatocele | 5 | 22.05 | 23.08 | 13.03 | 7.27 | 25.45 | 3.86 | 60.62 |
| Pneumonia | 38 | 13.22 | 12.4 | 9.67 | 4.4 | 18.6 | 0.08 | 59.85 |
| Psychosis | 18 | 22.89 | 13.22 | 19.36 | 15.56 | 24.94 | 5.2 | 53.88 |
| Scoliosis | 14 | 24.9 | 12.5 | 20.76 | 16.74 | 25.12 | 12.31 | 58.75 |

**Figure 1.**
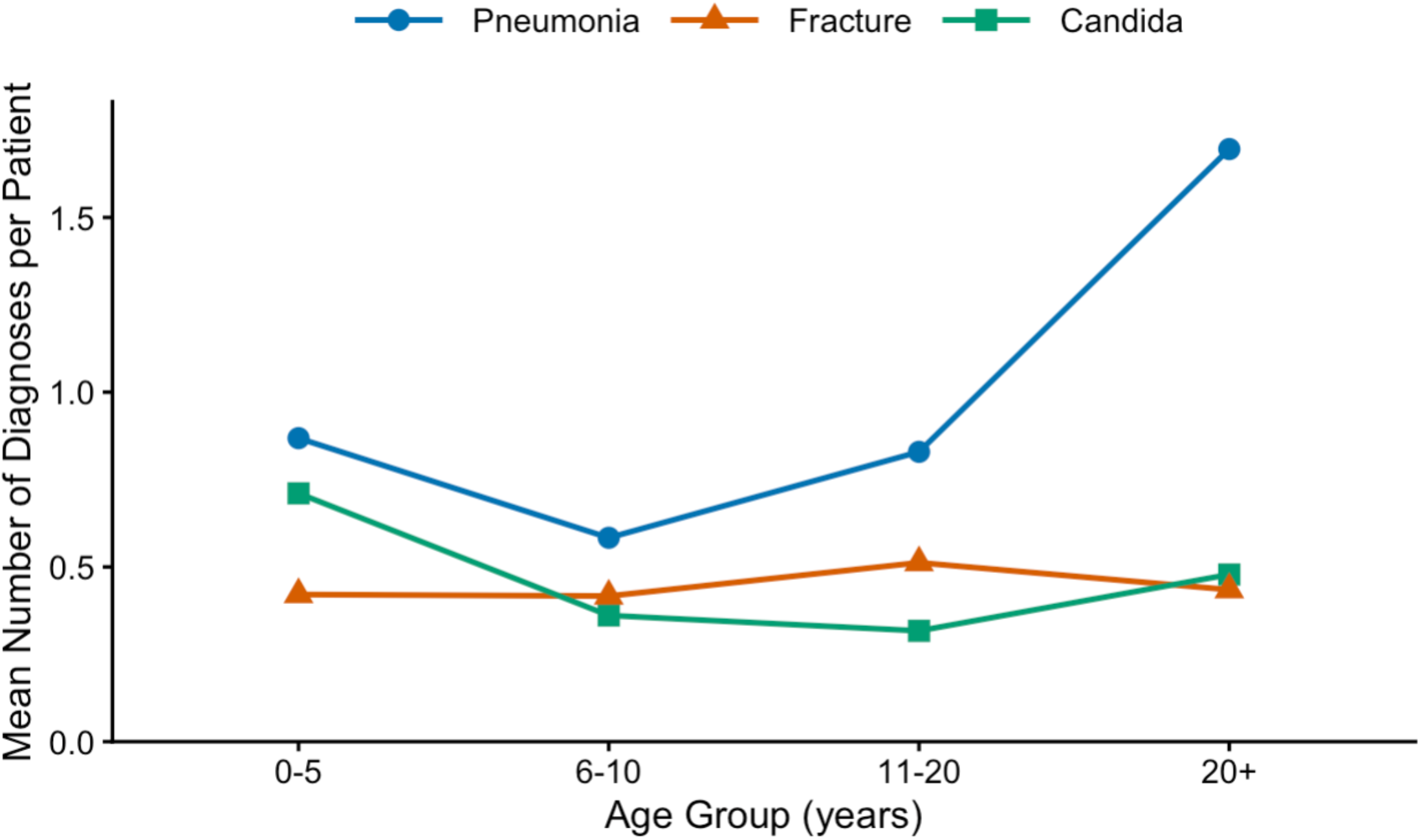

Although the number of adult patients in this cohort is relatively small compared to the pediatric patients, the frequency of pneumonia per patient is higher in the adult than in the children. This was not true for fractures or Candida.

## Discussion

Hyper IgE syndrome was one of the first inborn errors of immunity to be described with the classic descriptions of cold abscesses appearing in the 1960s ^2^. Nevertheless, it was many years before the genetic etiology was identified, and in spite of that the management approaches are still unclear ^14^. While various approaches have been promoted, the slow evolution of complications has hindered delineation of their impact. Most children with hyper IgE syndrome have significant infections that are problematic but treatable ^15^. Adults are known to develop complications related to pneumatoceles and aneurysms ^11^. But publications on the syndrome have been largely cross-sectional in nature. This study set out to better understand the trajectory of complications. As part of our initial data analysis, we extracted the most common complications from the entire cohort. To our surprise, developmental delay, and psychosis, which have not been previously shown to be associated with hyper IgE syndrome, affected a significant number of patients. Psychosis was found in 21% and developmental delay in 18% of the patients. STAT3 is widely expressed in non-neuronal cells in the brain. In fact, the protein expression in the brain is comparable to that seen in the connective tissues and bone marrow. Thus, there is a plausible mechanism of action to connect these observations to a cellular pathway.

The other main finding from this study is the increasing likelihood of pneumonia with age. Although for each patient, the constellation of complications can be very different and some patients had no pneumonia in adulthood, for some patients, the frequency of pneumonia was extremely high. With each pneumonia, the likelihood of pneumatoceles rises and pneumatoceles represent a significant risk factor for aspergillus. Only 9% of the patient cohort had aspergillus recorded as a complication, and only 6% had pneumatoceles recorder as a complication, suggesting that these feared disorders are not common, at least in the USIDNET registry.

There are limitations to the study. All patients in the USIDNET registry have been diagnosed by an immunologist which provides some assurance that the diagnosis is accurate. Nevertheless, not all patients had a genetic etiology recorded and this therefore represents a heterogeneous cohort. Whether these findings would be true across all etiologies is not known. The USIDNET registry records complications as ICD codes or text within the problem list. Some complications are not well represented by ICD codes and this is a significant limitation to this approach. In spite of that, this study constitutes significant new information that adds to our understanding about the evolution of features in hyper IgE syndrome. There is still much to learn regarding management, however, as a first step, identification of the features over time is an important first step.

## Data Availability

All data produced in the present study are available upon reasonable request to the authors

